# Evaluating the impact on clinical task efficiency of a natural language processing algorithm for searching medical documents: Prospective crossover study

**DOI:** 10.1101/2022.05.24.22275490

**Authors:** Eunsoo H Park, Hannah I Watson, Felicity V Mehendale, Alison Q O’Neil

## Abstract

**Background:** Information retrieval (IR) from the free text within Electronic Health Records (EHRs) is time-consuming and complex. We hypothesise that Natural Language Processing (NLP)-enhanced search functionality for EHRs can make clinical workflows more efficient and reduce cognitive load for clinicians.

**Objective:** To evaluate the efficacy of three levels of search functionality (no search, string search, and NLP-enhanced search) in supporting IR for clinical users from the free text of EHR documents in a simulated clinical environment.

**Methods:** A clinical environment was simulated by uploading three sets of patient notes into an EHR research software application and presenting these alongside three corresponding IR tasks. Tasks contained a mixture of multiple choice and free text questions.

A prospective crossover study design was used, for which three groups of evaluators were recruited, comprised of doctors (n=19) and medical students (n=16). Evaluators performed the three tasks using each of the search functionalities in an order according to their randomly assigned group. The speed and accuracy of task completion was measured and analysed, and user perceptions of NLP-enhanced search were reviewed in a feedback survey.

**Results:** NLP-enhanced search facilitated significantly more accurate task completion than both string search (5.26%, p=0.01) and no search (7.44%, p=0.05). NLP-enhanced search and string search facilitated similar task speeds, both showing an increase in speed over no search function (15.9%/11.6%, p=0.05). 93% of evaluators agreed that NLP-enhanced search would make clinical workflows more efficient than string search, with qualitative feedback reporting that NLP-enhanced search reduced cognitive load.

**Conclusions:** To the best of our knowledge, this study is the largest evaluation to date of different search functionalities for supporting target clinical users in realistic clinical workflows, with a 3-way prospective crossover study design. NLP-enhanced search improved both accuracy and speed of clinical EHR IR tasks compared to browsing clinical notes without search. NLP-enhanced search improved accuracy and reduced the number of searches required for clinical EHR IR tasks compared to direct search term matching.

## Introduction

The benefits of the transition from storing patient information in paper notes to Electronic Health Records (EHRs) has been a topic of debate among healthcare professionals [1-4]. Many clinicians have expressed dissatisfaction with their current hospital systems and EHR use is consistently cited as a contributor to clinician burnout [5-7]. Approximately 40% of doctors’ time is spent documenting patient information, with evidence showing that this work burden has increased following EHR implementation [8,9]. However, difficulties in quickly and accurately retrieving relevant information from these documents means this wealth of collected information is often not fully utilised [10,11]. Navigating EHR documents is challenging due to the complexity of medical text, which tends to include frequent misspellings, abbreviations, specialty-specific acronyms, and clinical shorthand [12-15]. Time-consuming and inaccurate information gathering from EHRs limits the efficiency of wider clinical workflows [16], with some doctors believing that difficulties in retrieving patient information significantly impact face-to-face patient care [17].

Despite the increasing sophistication of general search engines, there remain relatively limited search options within medical record software. One barrier is the need for patient data to be held securely; therefore, access to computing power and shared resources may be limited. To have clinical utility, search facilities must be fast and intuitive for use by time-pressured clinicians, including relatively junior members of staff to whom the task of searching through complex notes is frequently delegated. In addition, the search must handle high variability of text expression as mentioned above. Clinical text is error-prone; unlike journals and other publications, there is no editorial control to check for errors. Medical terminology, acronyms and abbreviations vary between regions, hospitals and even across different specialties; for instance, “CHD” may be related to chronic heart disease (Cardiology), congenital heart disease (Paediatrics), or congenital hip dislocation (Orthopaedics). Since clinical care is a high stakes environment, failure to find relevant information potentially has great implications; to effectively save the time of clinicians, search tools should ideally go beyond document-level results to locate and highlight all relevant sentences or even words within a document. Efforts to achieve easier information retrieval (IR) have included the integration of string search in some EHRs, similar to the “Ctrl-F” or “Find” function now frequently available on everyday platforms [18]. However, the effectiveness of string search is limited for heterogeneous clinical text; therefore, studies have also considered semantic search algorithms [19-22]. A large-scale retrospective analysis of searches performed in an EHR found that the use of search varied considerably across and within user roles, with physicians and pharmacists being the most active user groups [19]. A review of the use of search within EHRs found that few articles focussed on the impact of search within clinical workflows [23]; one study with 7 diabetes experts found that content-based search was both faster and more accurate than conventional search for finding relevant information [20], another study with 10 family and internal medicine physicians found that semantic search gave faster medical notes navigation for IR tasks [21], and a final study with 4 students found that a semantic search tool enabled faster clinical note summarisation [22]. Only one of the described studies [20] used a crossover study design. In this paper, a larger study is reported (n=35 valid task completions, n=42 qualitative responses), in which a 3-way prospective crossover study was conducted, comparing standard string search with no search and with Natural Language Processing (NLP)-enhanced search. The custom NLP-enhanced search tool combines ontologies with fuzzy matching to offer search functionality which captures not only semantically related terms (e.g., synonyms and hyponyms) but also linguistic alternative (miss)spellings and word forms of the search term. A simulated clinical environment was used alongside target-user feedback to determine whether search tools could make clinical workflows more efficient and reduce clinicians’ cognitive burdens when attempting to find information.

### 1.2 Aims and Hypotheses

#### Aims

To quantitatively and qualitatively compare the efficacy of three search functionalities for IR from medical free text documents, in terms of accuracy, speed, and ease of search.

#### Hypotheses

Search tools will allow clinical users to perform simulated clinical IR tasks faster and more accurately than when using no search, with the use of NLP techniques enabling NLP-enhanced search to perform more effectively than string search.

## 2 Materials and Methods

### 2.1 Search tools

The string search function is an open-source JavaScript library implementation^a^. NLP-enhanced search is a proprietary rule-based algorithm (developed at Canon Medical Research Europe) that leverages NLP techniques such as edit distance and stemming in conjunction with medical knowledge bases, notably the Unified Medical Language System (UMLS) semantic web, Metathesaurus [24], and the Wikipedia [25] and OpenMD medical abbreviation lists [26]. These sources are used to expand the original search term into a list of equivalent terms which are then located in the text. The tool was designed to locate linguistic variants such as misspellings and alternative spellings, wordforms, and abbreviations, as well as additional semantic synonyms.

Search tools were integrated into a patient-centric viewer (EHR research software), which allowed the user to type in a search term and view the highlighted findings within the retrieved subset of documents, which the user could scroll through. In the case of no search, the user was expected to scroll through the patient’s EHR to find the relevant information. Figure 1 illustrates the difference between the two search tools in the patient-centric viewer.

**Figure 1:**
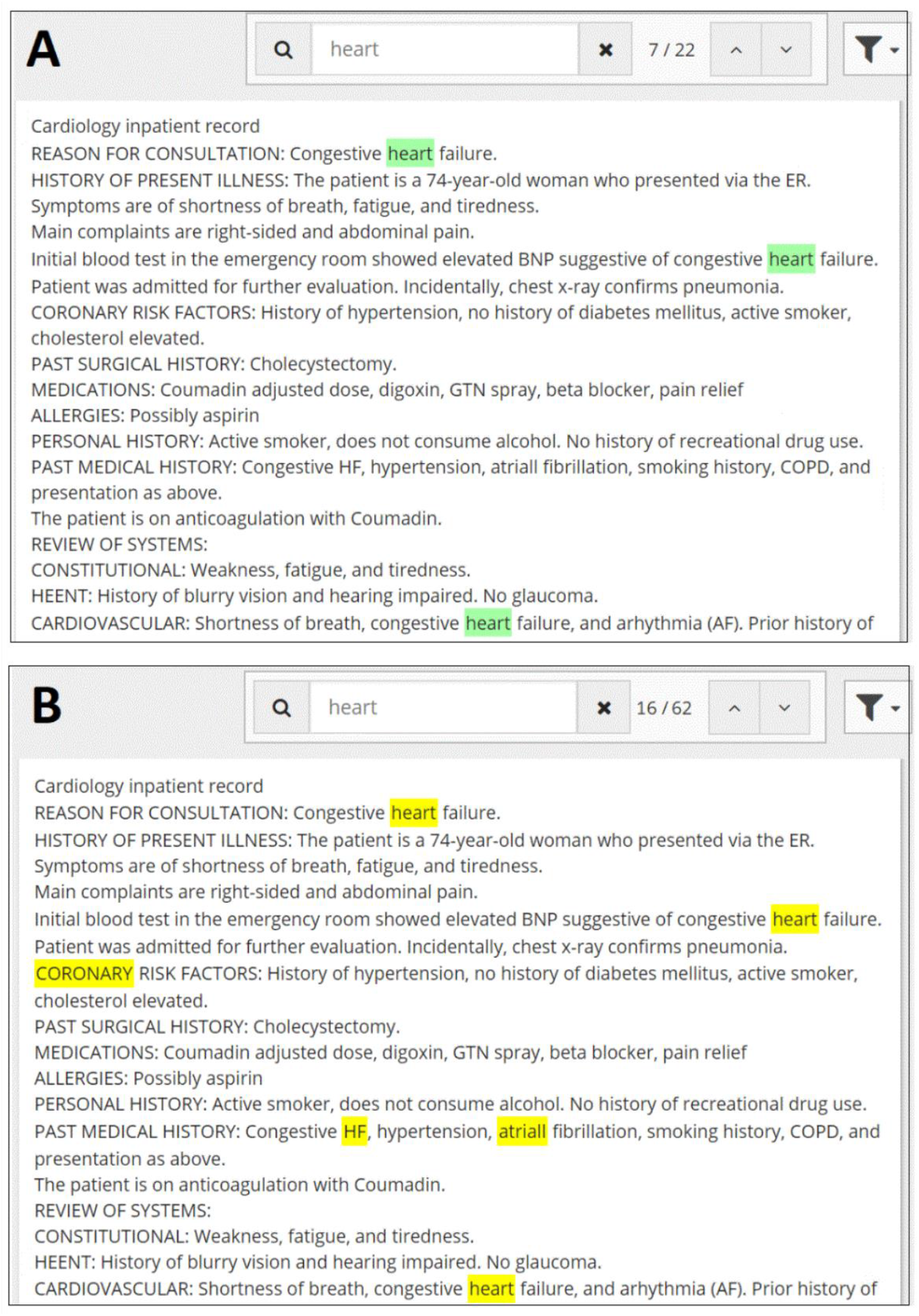
Example results for string search (A) and NLP-enhanced search (B) for the search term “heart”. String search returns only direct matches to “heart” (green highlights) whereas NLP-enhanced search returns also semantically related terms (yellow highlights), such as: “coronary”, the misspelling of atrial (fibrillation) as “atriall”, and the appearance of “heart” within the abbreviation of heart failure, “HF”.

### 2.2 Simulating a Clinical Workflow

Free text medical documents were synthesised for three fictional patients. These materials were paired with corresponding sets of ten IR questions for each patient, grounded in relevant and realistic clinical scenarios. Patient documents were uploaded into the patient-centric viewer. Questions were uploaded onto a custom Evaluation Platform built using Vue.js that also displayed the clinical scenarios and task-specific instructions for the evaluator. Below we describe the document synthesis and question generation in more detail.

#### Patient document synthesis

Three patient profiles were created with varying age, sex, ethnic background, social history, and medical history. The three patients were assigned primary medical specialties of respiratory, neurology, and oncology. For each patient, 20 documents were created by selecting and augmenting publicly available anonymised medical documents [27], as well as manually synthesising additional documents, to provide a patient EHR with a coherent chronological sequence of clinical events. Documents were varied and included discharge letters, outpatient clinic letters, operation notes, and general practice (GP) referral letters. To imitate real-world medical text, common misspellings, abbreviations and acronyms were included in the text, using investigator clinical experience (HW) and reference papers [13].

#### Clinical scenarios and question generation

For each task, clinical scenarios were designed to focus on real-world situations where information can be extracted from patient notes. To ensure the tasks were comparable across patients (and therefore interventions), a master template of 10 questions prompting IR was created, which was then tailored to fit each patient scenario. Questions were inspired by past medical exam questions [28] and investigators’ clinical experience (HW, FM). Requested information resembled that required in typical clinical workflows to support clinical decision-making. Care was taken to ensure task questions tested search function and not clinical knowledge or judgement, therefore all answers could be found by searching the respective patient’s notes. Questions required a mixture of multiple choice and free text responses. Examples of scenarios and corresponding questions for each patient can be seen in Figure 2.

**Figure 2:**
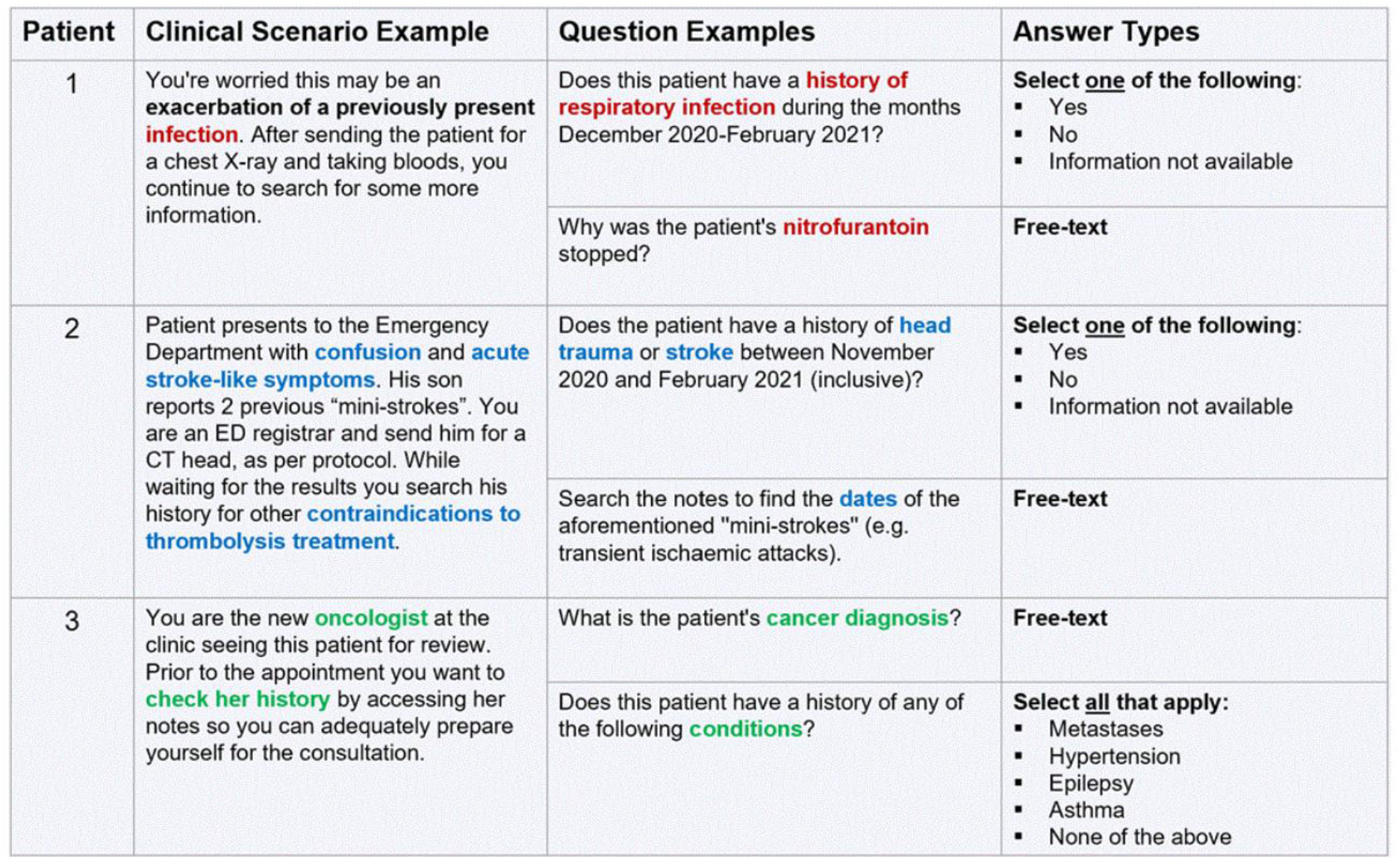
Examples of clinical scenarios for each patient and their corresponding question-answer options. Scenarios aimed to simulate a standard clinical workflow, providing context for the questions.

### 2.3 Study Design

The clinical evaluation pipeline was structured as a prospective crossover trial design. Evaluators were banded based on their level of clinical experience before being assigned pseudonymised evaluator IDs that were used for the remainder of the study and analysis. Evaluators in each band were then randomly allocated across the three study groups using a random number generator. This yielded three groups stratified for level of clinical experience. Each group had a predetermined order of search functionality; once the three tasks had been completed using the allocated search order, evaluators were asked to fill out a feedback survey that focused on their user experience.

### 2.4 Evaluator Recruitment and Training

Recruitment for the study was accomplished via professional contacts and advertising on social media channels to reach evaluators from a variety of clinical specialties and years of clinical experience.

A training video was provided to evaluators, which comprised a brief introduction to the study, demonstrations of the three search interventions within the patient-centric viewer and detailed instructions on how to complete the evaluation. An example patient with a small set of curated medical documents was also provided for training, on which evaluators could familiarise themselves with the capabilities of the different search functionalities.

### 2.5 Data Collection

Evaluators were provided with secure remote access to the evaluation environment (Figure 3), allowing the evaluation to be performed remotely from personal machines. Using this setup, evaluators could view the patient-centric viewer and the Evaluation Platform. Answers had to be inputted sequentially on the Evaluation Platform, which did not allow evaluators to return to a question once they had submitted an answer.

**Figure 3:**
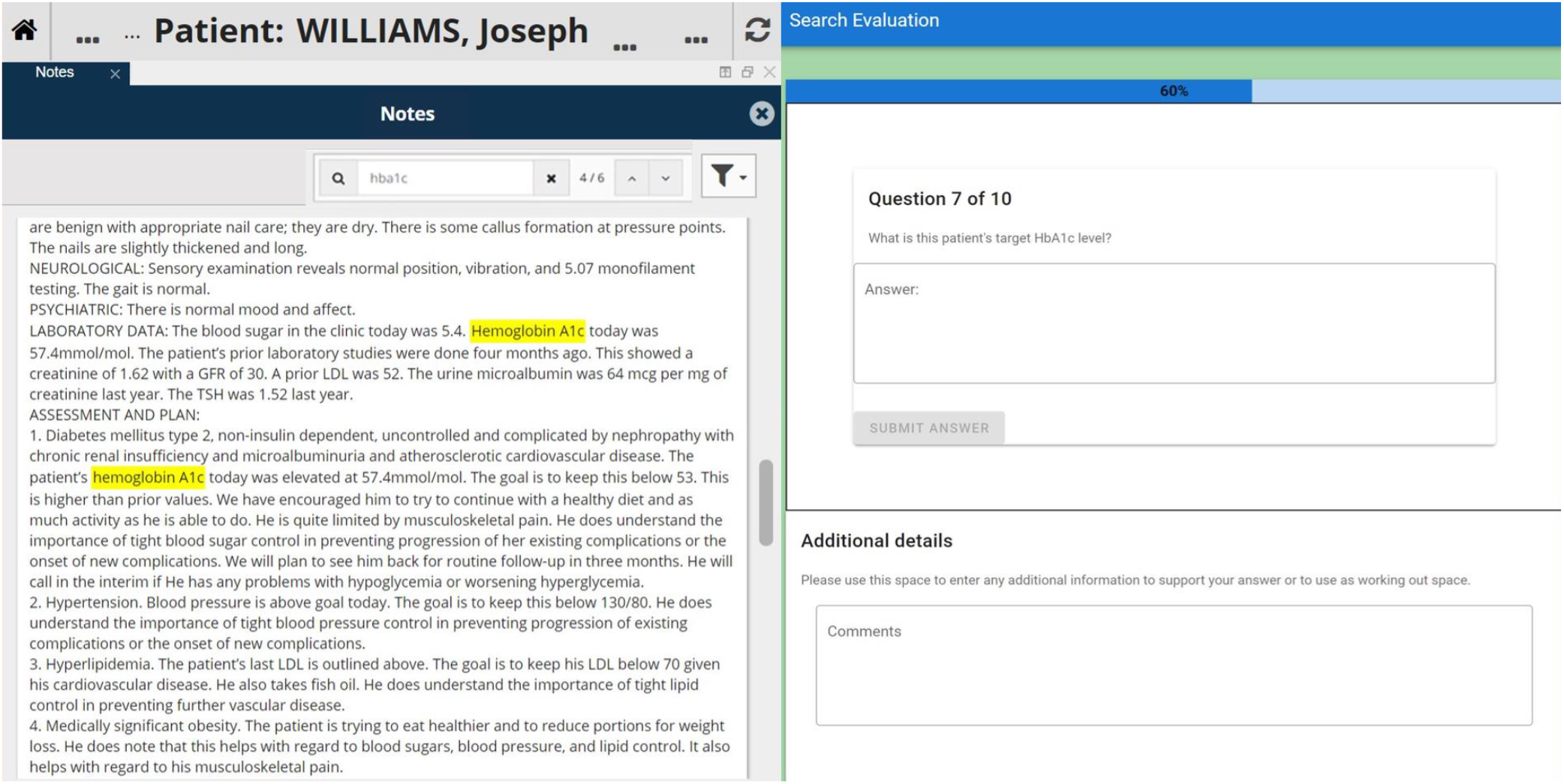
Screenshot of evaluation environment during a task. Evaluators only had permission to view the two relevant sites: the patient-centric viewer (left) and Evaluation Platform (right). The patient-centric viewer contains the synthetic patient documents for a given patient (in this case “Joseph Williams”) with “hba1c” as the search term. The Evaluation Platform detailed the clinical scenarios, task-specific instructions, along with the question-and-answer sections.

During each task, the evaluators submitted answers to the task questions through the Evaluation Platform. To ensure accurate recording of task times, evaluators were asked to do each task in one go, and to take breaks between tasks rather than during tasks. Evaluators were free to spend as long as they needed on each task. In addition, a search log was kept that recorded the search terms entered by the evaluator along with the search functionality used, plus the time spent on each question.

### 2.6 Data Analysis

#### Exclusion criteria

Data was excluded where search logs showed that evaluators had used an incorrect search functionality for a given task.

#### Question marking

Two clinical investigators (EP, HW) came to a consensus on the correct answers for each question. Answers were then clustered depending on the document in which they were located, and marks were awarded for finding each relevant area of correct documents. For example, if 3 pieces of clinical information across 2 unique documents were required to correctly answer a question, then 2 marks were awarded if the correct answer was inputted as the evaluator had successfully found both documents. Questions were weighted equally.

#### Statistical analysis

Data analysis was performed using custom Python code. For all metrics, samples were weighted to compensate for imbalances in group size (see Section 3.1). Paired two-tailed t-tests were performed to determine if there was a significant difference in timing and accuracy between 1) string search and no search, 2) NLP-enhanced search and no search, and 3) NLP-enhanced search and string search.

#### Search term analysis

Following the study, search term logs were analysed to extract the number and pattern of search terms for each type of search.

## 3 Results

### 3.1 Evaluator Demographics and Group Stratification

In total, 60 evaluators were recruited with multiple levels of clinical experience from medical students to doctors, and from 9 specialties ranging from vascular surgery to general practice. Of 60 recruited evaluators, 44 evaluators completed the tasks; 35 were included in the final analysis (Table 1), whilst 9 were excluded. Evaluators were excluded from the quantitative analysis if their data was corrupted (n = 2) or they completed the tasks incorrectly (n = 7), for example by using the wrong search functionality for a given task. From the original 20 evaluators per group, this gave n = 7 (Group 1), n = 13 (Group 2) and n = 15 (Group 3) successful completions. There were 42 responses to the feedback survey. Table 1 shows the final distribution of clinical experience across the groups.

**Table 1:** Summary table showing the allocation across clinical bands and study groups.

| Clinical band | Group 1 | Group 2 | Group 3 | Total |
| --- | --- | --- | --- | --- |
| Medical students |  |  |  |  |
| • Pre-clinical (years 1 – 3) | 4 | 3 | 3 | **10** |
| • Clinical (years 4 – 6) | 0 | 4 | 2 | **6** |
| Doctors |  |  |  |  |
| • 1 – 5 years clinical experience | 0 | 3 | 3 | 6 |
| • 6 – 10 years clinical experience | 1 | 1 | 4 | 6 |
| • 11+ years clinical experience | 2 | 2 | 3 | 7 |
| **Total** | **7** | **13** | **15** | **35** |

### 3.2 Effect of search functionality on speed and accuracy of task completion

Results are shown in Tables 2 and 3. Overall, NLP-enhanced search facilitated significantly more accurate task completion than both string search (p=0.01) and no search (p=0.05). In terms of speed, NLP-enhanced search and string search gave significantly faster task completion compared to no search (p=0.05); there was no significant time difference between string search and NLP-enhanced search.

**Table 2:**
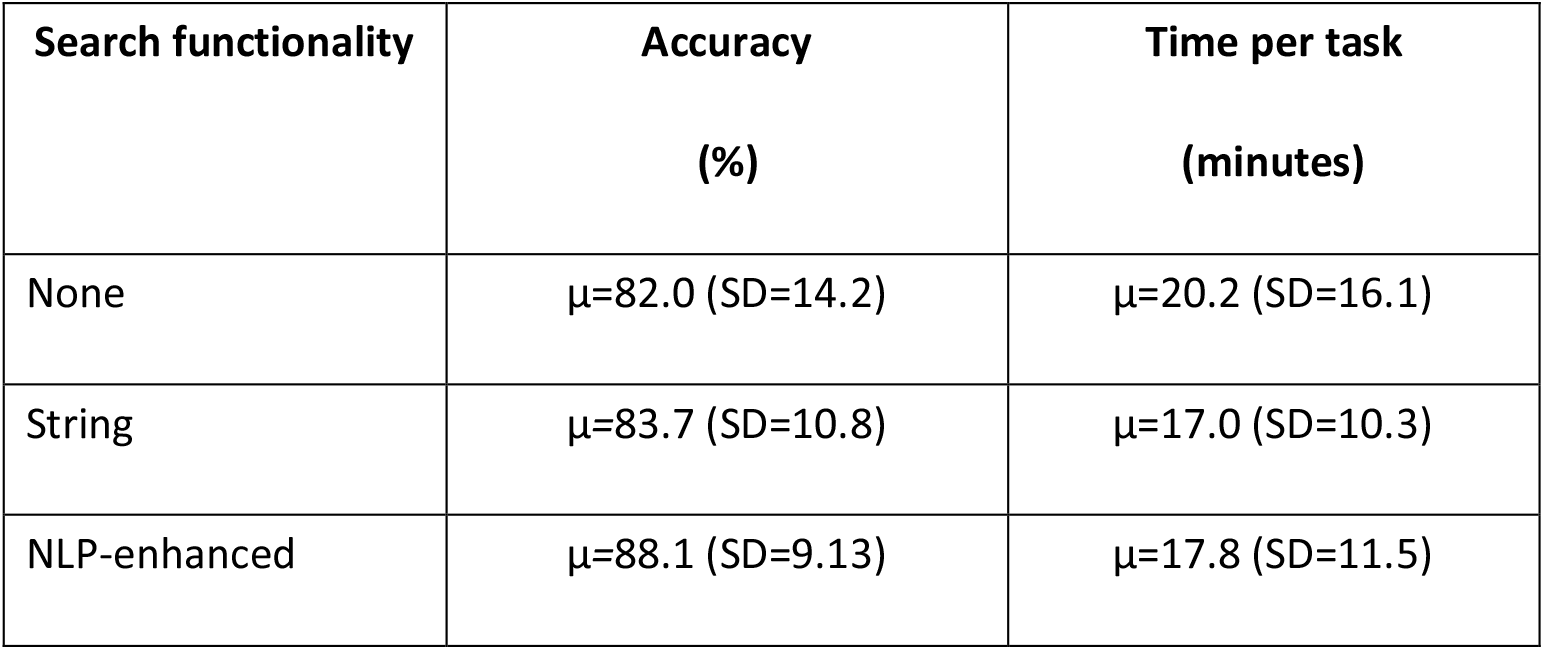
Accuracy and time for different search functionalities, showing µ=mean (SD=standard deviation).

**Table 3:**
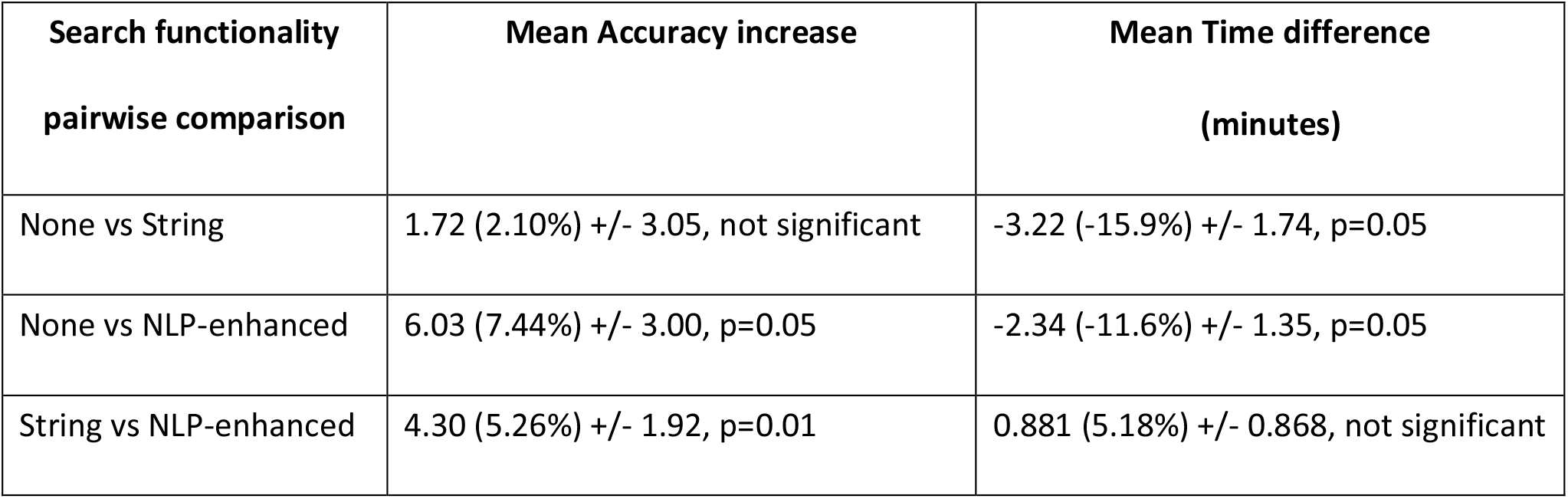
Pairwise comparison between different search functionalities, showing mean difference +/- standard error and p-value significance level.

### 3.3 Analysis of search terms employed by evaluators

Analysis of the logged search terms (Table 4) revealed that evaluators tried almost twice as many search terms when using string search compared to NLP-enhanced search, and uptake of string search was slightly lower than NLP-enhanced search i.e., the percentage of questions for which no searches were performed was higher for string search.

**Table 4:** Analysis of used search terms, showing: the percentage of answers that used search, and the number of search terms for each of these answers i.e., µ=mean (SD=standard deviation). Best results shown in bold.

|  | Answers using Search<br>(%) | Number of Search Terms per Answer |
| --- | --- | --- |
| String Search | 83.7 | 3.51 (SD = 2.91) |
| NLP-enhanced Search | **95.1** | **2.05 (SD = 1.49)** |

The higher number of search terms required for string search might intuitively be explained by the user needing to try multiple synonyms to find relevant information. For instance, for the question “*Does the patient have a history of stroke?*” in the text there were 4 negative mentions scattered through the documents: “does not look like she has a stroke”, “No TIA or CVA”, “No CVA” and “No CVA” (TIA = transient ischaemic attack; CVA = cerebrovascular accident). NLP-enhanced search found all mentions with the search term “stroke” (which was the only term that evaluators tried), but string search evaluators also tried “TIA”, “CVA”, “neurological”, “history” and “infarction” in their efforts to find all relevant information. Interestingly, we see that evaluators were sometimes searching for neighbouring words (“history”, “neurological”) most likely as a method to bypass the possible variation in textual mentions. Further, string search does not match spelling variants (or misspellings) and therefore evaluators sometimes tried different spellings e.g., for the question “*Is the patient currently on full-dose anticoagulant treatment?”*, both “anti-coagulant” and “anticoagulant” were tried as successive search terms by evaluators using string search.

This analysis also highlighted that the strict parameter settings for string search meant that search terms matched only to whole words, not to substrings and thus evaluators could not search with a prefix. We see some evidence of evaluators adjusting to this e.g., searching first for “anticoag” and then “anticoagulant”, or searching for both “smoke” and “smoker”, and this also increases the number of search terms tried.

### 3.4 User perceptions of NLP-enhanced search

User perceptions were assessed via the feedback survey which included a mix of Likert scale ratings, from “strongly disagree” to “strongly agree”, and free text responses. Below we summarise responses to four questions.

#### How was NLP-enhanced search perceived?

Most respondents positively described the capabilities of NLP-enhanced search, noting its identification of misspellings, wordforms and synonyms, though some reported that NLP-enhanced search returned too many findings (*“[NLP-enhanced] search was very clever and thorough but could return 100 results”)*. However, when rating the efficacy of NLP-enhanced search, 76% of respondents thought any unrelated findings, i.e., *false positives*, did not significantly impact the usefulness of the search algorithm.

#### Is NLP-enhanced search better than string search?

81% of respondents agreed that NLP-enhanced search facilitated more relevant IR than string search. However, many commented that the string search capabilities within the patient-centric viewer were more limited than they were accustomed to on everyday devices, stating that “*string search was too discriminatory*” (the parameter settings meant that only whole word matches were returned, not substrings, as discussed in Section 3.3).

#### Would NLP-enhanced search make clinical workflows more efficient?

93% (39 out of 42) respondents agreed that NLP-enhanced search would make clinical workflows more efficient than string search, in particular during clinics and clerking of patients. Free text feedback reflected this, with respondents reporting that NLP-enhanced search was useful and less time-consuming when compared to string search or no search when retrieving specific information. One evaluator commented *“the [NLP-enhanced] search tool made it significantly easier for me to find the information I was looking for and also quicker”*. On the other hand, respondents further reported that NLP-enhanced search would not always be the best method for situations where a comprehensive overview of a patient is needed. In this case, assimilating information using manual review (no search) would be more effective. One evaluator said, *“I felt that using the [NLP-enhanced] search tool meant I wasn’t focussing on the case as much but just looking for words”*. A common opinion was that NLP-enhanced search would be a useful addition to manual review for clinical tasks.

#### Would NLP-enhanced search reduce cognitive load?

Respondents frequently mentioned that NLP-enhanced search made it easier to retrieve the information they were looking for, with one evaluator stating that *“[NLP-enhanced] search is an excellent tool for a quick way to filter through relevant information”*. While a few mentioned that too many results were returned, respondents also reported that going through the relevant findings was easier and preferable to a full manual review of the notes, with manual review being described as “*tedious”, “painstaking”* and *“very easy to miss vital information”*. One evaluator commented that NLP-enhanced search could *“improve the workload of an already overworked profession”*.

## 4 Discussion

Our results showed a significant increase in accuracy when NLP-enhanced search was used compared to string search and no search, whilst both NLP-enhanced search and string search offered time savings. There was a perception of easier navigation from evaluators and a measured decrease in required interactivity in the case of NLP-enhanced search (lower number of search terms compared to string search). We caveat this conclusion with the observation that the strict parameter settings of string search meant search terms matched only to whole words, not substrings; this increased the number of terms that evaluators used and potentially reduced the search accuracy, compared to a string search version that matches also to substrings.

There is limited literature on the potential impact of EHR search tools on day-to-day clinical care [29]. Our results support those of previous studies [20-22] which find that semantic search tools allow faster and more accurate EHR task completion in simulated clinical workflows. A related study showed Artificial Intelligence-optimised patient records to give speed improvements in answering clinical questions whilst maintaining the same accuracy [30]. Interestingly, the impact of the patient record search engine MorphoSaurus has been measured in a real clinical setting [31], albeit with user surveys only. This method would have had the benefit of involving real-world stresses such as task interruptions and time pressure, as well as the key element of patient interaction. Importantly, however, our method of using a controlled simulated clinical environment enabled us to control for variables such as distractions or interruptions, as well as variation in the complexity and length of medical records. Additionally, our crossover design controlled for individual participants’ ability, experience and diligence. This enabled robust comparison of quantitative and qualitative data for each search type, while minimising the impact of confounding factors.

Overall, evaluator feedback suggested that the optimum approach to navigating clinical notes is a hybrid of manual browsing and search, depending on the context. In the real-world, NLP-enhanced search is likely best employed as a complementary tool to aid clinical users in navigating clinical notes, with the ability to manually parse and ingest relevant facts from a complex medical history remaining important.

## 5 Conclusion

In conclusion, this study suggests that search tools have a positive effect on both the measured and perceived accuracy and ease of clinical IR. Search tools that can leverage NLP techniques are more effective for retrieving all relevant terms from heterogeneous medical free text. There is potential to reduce clinicians’ cognitive burden and make clinical workflows more efficient. A critical direction for future research is to assess the use of search tools in real-world clinical practice.

## Data Availability

All data used is non-publicly available data.

## Authors’ Contributions

- **Eunsoo H Park** co-designed the study, co-designed the patient histories, reviewed the synthetic patient notes, designed the tasks, designed the clinical feedback survey, organised evaluator recruitment, recorded training materials for evaluators, performed preliminary analysis of the findings, and contributed to the paper draft.
- **Hannah I Watson** co-designed the study, co-designed the patient histories, created the synthetic patient notes, reviewed the tasks, reviewed the clinical feedback survey, supported evaluator recruitment, organised the infrastructure for the practical evaluation, contributed to & reviewed the analysis, and contributed to & reviewed the paper draft.
- **Felicity V Mehendale** co-designed the study, reviewed the patient histories, reviewed the synthetic patient notes, reviewed the tasks, reviewed the clinical feedback survey, reviewed the analysis, and contributed to & reviewed the paper draft.
- **Alison Q O’Neil** co-designed the study, organised provision of the NLP-enhanced search, reviewed the tasks, reviewed the clinical feedback survey, contributed to & reviewed the analysis, and contributed to & reviewed the paper draft.

## Acknowledgements

Thank you to Professor Keith W Muir (Institute of Neuroscience & Psychology, University of Glasgow) for his clinical insights during the development of the NLP-enhanced search tool. We would like to thank the West of Scotland Safe Haven within NHS Greater Glasgow and Clyde for assistance in creating and providing a dataset which was used during development of the NLP-enhanced search tool.

Many thanks to the Canon Medical Research Europe staff who developed the infrastructure required for this evaluation: Yvonne Belton, Michael Corrigan, Vismantas Dilys, Francisco Gomez, Graham Jones, Hamish MacKinnon, David Miller, Emel Muzaç, Paul Norman, and Euan Robertson. Further, we would like to acknowledge the research team which was responsible for creating the NLP-enhanced search tool: Murray Cutforth, Vismantas Dilys, Matúš Falis, Aneta Lisowska, Hamish MacKinnon, Maciej Pajak, Alison O’Neil, and Hannah Watson.

Thank you to our evaluators: Fiona Auld, Anna Barton, Rong Bing, Cameron Brown, Khai Syuen Chew, Jane Yi Chiam, Vanessa Chou, Luisa Ciriello, George Cooper, Iona Cutworth, Jamie Donachie, Vivienne Evans, Magdalena Gabrysiak, Eilidh Gunn, Mohamed Hamed, Hamzah Hanif, Ewen Harrison, Kylla Hernandez, Lana Huang, Katie Hunter, Haider Khan, David Kluth, Niki Kouvroukoglou, Barbora Krivankova, Tommy Le, Charles Leeson-Payne, Alinah Sum-Ping Leung, Jenny Lockhart, Jack Lugue, Angus MacLeod, Tomos Morgan, Ellen Murgitroyd, Sarah Murphy, Helen O’Neil, Yusuke Onishi, Lisa Ragunathan, Nikita Rana, Qi Shun Yong, Lucy Taylor, Evangelos Tzolos, Miriam Veenhuizen, Philippa Veenhuizen, Olivia Yu, Sydney Zides.

Thank you to our pre-trial evaluators: Marcus Boyd, Elizabeth Daly, Greta Economides, Keziah Lewis, Abhishek Nambiar, Sumrah Naqvi, Risako Sakatsume, Faye Sikora, Emma Warburton.

Thank you to our internal Canon reviewers: William Clackett, Russell Hung and David Miller.

Many thanks to MTSamples for permitting free use and modification of their data to create the patient case studies.

## Abbreviations

EHR: Electronic Health Record
IR: Information Retrieval
NLP: Natural Language Processing
UMLS: Unified Medical Language System

## Footnotes

a https://markjs.io/

